# Presurgical immune biomarkers associated with pain intensity and pain interference recovery after total knee arthroplasty: findings from the PRIME-KNEE study

**DOI:** 10.64898/2026.06.15.26355689

**Authors:** Corey B Simon, Virginia B Kraus, Janet L Huebner, Marissa C. Ashner, Akshay Bareja, Sarah Peskoe, Katherine S. Hall, Heather E. Whitson, Cathleen S. Colón-Emeric

## Abstract

Chronic postsurgical pain (CPSP) prevalence after total knee arthroplasty (TKA) is >20%. Circulating immune biomarkers are associated with musculoskeletal pain but poorly understood as CPSP predictors. This prospective, longitudinal study of 203 patients undergoing TKA tested presurgical plasma biomarkers associated with 6-month CPSP, using approaches from geriatrics biomarker research: expected recovery differential (ERD; resilience outcome) and penalized, machine-learning regularization modeling (elastic net and LASSO regression). Forty-nine presurgical candidate biomarkers were considered. CPSP was operationalized using ERDs built around PROMIS pain intensity and pain interference, which quantified the difference between observed and expected recovery after accounting for demographic, comorbidity, reserve, and perioperative factors. Plasma/ERDs from ∼130 patients revealed 13 biomarkers with the highest selection stability criteria, and either positive or negative (+/–) associations with ERDs. Interleukin (IL) −5 (–) and Lipopolysaccharide-Binding Protein (LBP; +) were associated with both ERDs. Unique associations with pain intensity ERD included Cytomegalovirus-Specific IgG Negative (CMV IgG–; –), Macrophage Inflammatory Protein-1 Beta (MIP-1β; –), IL-12p70 (–), Cluster of Differentiation 30 (sCD30; –), Interferon alpha 2a (IFNα2a; +), and Leukemia Inhibitory Factor (LIF; +). Unique associations with pain interference ERD included Lipopolysaccharide (LPS; –), Activin A (–), IL-8 (–), Serum Amyloid A (SAA; –), and IL-7 (+). Protein-protein interaction and topological analyses suggest a centralized subnetwork with higher-than-expected connectivity involving IL-5, IL-7, IL-8, MIP-1β, and IFNα-2a, among others. This study proposes rigorous yet feasible approaches to expedite pain biomarker research and introduces presurgical biomarkers to consider in future TKA-CPSP biosignature derivation.

**SUMMARY:** Methodological approaches used in geriatric biomarker studies were applied to identify 13 pre-surgical plasma biomarkers associated with chronic postsurgical pain outcomes following total knee arthroplasty.

## 1. INTRODUCTION

Knee osteoarthritis (OA) is highly prevalent among older adults and associated with chronic pain and disability. [4,5,20,35]. While total knee arthroplasty (TKA) is effective for many, more than 20% of patients experience chronic postsurgical pain (CPSP) [67]. CPSP is pain persisting ≥ 3 months after surgery [107,113], and is heavily influenced by presurgical self-reported pain intensity and cognitive/affective factors [70,87,88]. However, self-report measures are not biomarkers (“*a biomarker is not an assessment of how an individual feels, functions, or survives* [29,96]”), which limits the utility of such measures in clinically-meaningful risk stratification or treatment tailoring [21,22,29,45,74,98,106,114].

Circulating immune biomarkers (e.g., C-reactive protein) have an established association with musculoskeletal pain outcomes [8,28,30,53,61,81,110], and are a cornerstone of precision health-focused pain research [22,26,27,69,112]. As CPSP predictors, however, immune biomarkers are poorly understood [75], and a major reason why is patient heterogeneity. Patients who undergo TKA for knee OA pain have a wide range of demographic, biological, health, surgical, and pain-related factors [62,68,134]. If these factors are not accounted for, then isolating the predictive influence of CPSP biomarkers will be difficult [7,96,112,124]. Unfortunately, traditional methods make rigorous covariance control both time and resource-intensive, which means a large proportion of studies today have a high risk of bias or a low probability of replication.

Facing similar barriers, researchers in aging and geriatric medicine have prioritized alternative approaches to biomarker discovery that include novel outcomes and modeling [33,44,55,60,95,136]. For example, ‘expected recovery differential’ (ERD), which is a resilience outcome that quantifies better-than-expected versus worse-than-expected recovery after accounting for perioperative factors [1,17,18,128]. Penalized, machine-learning regularization (e.g., LASSO [38]) is a mathematical and statistical approach to isolate biomarkers from confounders, as well as other biomarkers [51,64,135]. In a previous study of older adults with hip fracture, our team combined ERD with LASSO to identify several age-related biomarkers that explained >25% of the variance in recovery [94].

The current study is a planned extension of past work to investigate presurgical plasma biomarkers associated with CPSP ERDs 6 months after TKA. We derived separate postsurgical ERDs for pain intensity and pain interference, based on guideline-based recommendations for CPSP assessment and growing recognition that pain interference is central to chronic pain, disability, health, and mortality risks [32,46,102,107,111,113,132,133]. Our primary aim was to identify a stable set of presurgical plasma biomarkers associated with CPSP ERDs. Our secondary aim was to use protein-protein interactions (PPI [116]) and topological analysis [16,86,125] to visualize plausible biological networks underlying CPSP ERDs. The overarching goal of this study was to integrate approaches from pain and geriatric research fields to generate mechanistic hypotheses for future CPSP biosignature derivation.

## 2. METHODS

### 2.1 Study Summary

#### Physical Resilience

*Indicators and Mechanisms in the Elderly* (PRIME-Knee) is a single-site, prospective cohort study designed to identify provocative tests and presurgical biomarkers that predict resilience to TKA [19,127]. Participants were 203 patients who underwent unilateral TKA at Duke University Medical Center. Enrollment criteria were age ≥60 years; community dwelling; elective TKA lasting ≥2 hours in duration; access to a Bluetooth-enabled mobile device (for actigraphy data used in a separate analysis); and ability to speak and understand English. Exclusion criteria were an inability to ambulate independently with or without an assisted device; known dementia or a failed cognitive screen; being a correctional facility inmate; medical treatment for non-skin cancer in the last 12 months; or vision or hearing impairment that prevented reliable cognitive assessment or telephone interviews after best accommodation.

Participants completed a presurgical study visit within the 6-week period before their scheduled TKA. Data collection included blood draws for plasma biomarkers in addition to clinical tests and measures used to derive pain intensity and interference ERDs. These covariates (see **Supplemental Table 1** for full list and description) included self-reported demographics, comorbidities, reserve (physical, psychosocial, cognitive), and surgical factors [127]. Participants completed Patient-Reported Outcomes Measurement Information System (PROMIS) self-reported pain intensity and interference questionnaires prior to surgery and 6 months after surgery. All performance assessments and questionnaires were completed in the research facility while questionnaires were administered either on site or remotely at the participant’s discretion. Study procedures were approved by the Duke University Institutional Review Board and participants provided informed consent.

### 2.2 Candidate Biomarkers

The Molecular Measures Core (MMC) performed all immunoassays within the Duke Biomarkers Core Facility, housed in the Duke Molecular Physiology Institute. A total of 48 presurgical biomarkers were chosen *a priori* [127] as candidates, based on significance in a previous study of hip fracture patients and/or presence in Pillars of Aging (adaptation to stress, epigenetics, inflammation, macromolecular damage, metabolism, proteostasis, stem cells and regeneration) processes and pathways [54]. Biomarker abbreviations, names, descriptions, and baseline values are listed in **Table 1**.

**Table 1.** Candidate Biomarker Reference Table.

| **Symbol** | **Biomarker** | **Type** | **Function** | **Units** | **Baseline Median/IQR\*** |
| --- | --- | --- | --- | --- | --- |
| **activin-A** | Activin A | Cytokine | Cell regulation | pg/ml | 269.83 (212.65–331.63) |
| **CMV IgG** | Cytomegalovirus-Specific IgG | Antibody | Immune exposure | EU/ml | 445.62 (130.30–746.99) |
| **CMV IgG-** | Cytomegalovirus-Specific IgG Negative | Antibody | No immune exposure | EU/ml | n=52 (40.9%)** |
| **cortisol** | Cortisol | Hormone | Stress response | mcg/dL | 8.70 (6.69–11.03) |
| **CRP** | C-Reactive Protein | Protein | Inflammation | ng/ml | 1901.96 (940.34–4222.38) |
| **D-dimer** | D-dimer | Protein | Clot activity | ng/ml | 532.79 (384.27–747.46) |
| **eotaxin** | Eotaxin | Chemokine | Immune cell recruitment | pg/ml | 268.56 (217.36–323.08) |
| **fibrinogen** | Fibrinogen | Protein | Clot formation | mg/ml | 3.91 (3.45–4.49) |
| **GDF-15** | Growth Differentiation Factor 15 | Protein | Stress signaling | pg/ml | 1072.69 (817.79–1451.13) |
| **GP130** | Glycoprotein 130 | Protein | Immune signaling | ng/ml | 254.18 (232.67–284.33) |
| **GRO-α** | Growth-Regulated Oncogene Alpha | Chemokine | Immune cell recruitment | pg/ml | 73.00 (44.93–137.34) |
| **GZMB** | Granzyme B | Protease | Cell death | pg/ml | 2.54 (1.93–3.66) |
| **ICAM-1** | Intercellular Adhesion Molecule-1 | Protein | Immune cell adhesion | ng/ml | 498.38 (435.27–574.01) |
| **IFN-α2a** | Interferon Alpha 2a | Cytokine | Antiviral activation | pg/ml | 1.06 (0.37–1.87) |
| **IFN-γ** | Interferon Gamma | Cytokine | Immune activation | pg/ml | 7.17 (4.63–10.59) |
| **IL-1β** | Interleukin-1 Beta | Cytokine | Inflammation | pg/ml | 0.05 (0.04–0.13) |
| **IL-1Ra** | Interleukin-1 Receptor Antagonist | Cytokine | Inflammation inhibition | pg/ml | 157.99 (121.90–230.45) |
| **IL-2** | Interleukin-2 | Cytokine | Immune activation | pg/ml | 0.29 (0.29–0.35) |
| **IL-4** | Interleukin-4 | Cytokine | Immune regulation | pg/ml | 0.01 (0.00–0.02) |
| **IL-5** | Interleukin-5 | Cytokine | Immune activation | pg/ml | 0.56 (0.31–0.82) |
| **IL-6** | Interleukin-6 | Cytokine | Inflammation | pg/ml | 1.39 (0.95–2.03) |
| **IL-7** | Interleukin-7 | Cytokine | Immune maintenance | pg/ml | 1.61 (0.84–2.34) |
| **IL-8** | Interleukin-8 | Chemokine | Immune cell recruitment | pg/ml | 7.44 (5.97–9.40) |
| **IL-10** | Interleukin-10 | Cytokine | Inflammation inhibition | pg/ml | 0.30 (0.20–0.39) |
| **IL-12p70** | Interleukin-12p70 | Cytokine | Immune activation | pg/ml | 0.23 (0.10–0.34) |
| **IL-13** | Interleukin-13 | Cytokine | Immune regulation | pg/ml | 0.33 (0.28–0.95) |
| **IL-15** | Interleukin-15 | Cytokine | Immune cell survival | pg/ml | 1.32 (1.17–1.55) |
| **IL-18** | Interleukin-18 | Cytokine | Immune activation | pg/ml | 724.01 (542.31–929.52) |
| **IL-27** | Interleukin-27 | Cytokine | Immune regulation | pg/ml | 128.32 (100.86–188.65) |
| **IL-37** | Interleukin-37 | Cytokine | Inflammation inhibition | pg/ml | 2.16 (0.34–5.64) |
| **IP-10** | Interferon gamma-inducible protein 10 | Chemokine | Immune cell recruitment | pg/ml | 510.25 (383.40–666.79) |
| **I-TAC** | Interferon-inducible T-cell Alpha Chemoattractant | Chemokine | Immune cell recruitment | pg/ml | 14.50 (10.59–19.30) |
| **LBP** | Lipopolysaccharide-Binding Protein | Protein | Immune activation | mcg/ml | 5.14 (3.63–6.12) |
| **leptin** | Leptin | Hormone | Energy signaling | ng/ml | 27.34 (15.21–49.86) |
| **LIF** | Leukemia Inhibitory Factor | Cytokine | Cell regulation | pg/ml | 2.00 (1.26–3.20) |
| **LPS** | Lipopolysaccharide | Glycolipid | Immune activation | EU/ml | 100.40 (80.24–124.60) |
| **MCP-1** | Monocyte Chemoattractant Protein-1 | Chemokine | Immune cell recruitment | pg/ml | 352.37 (310.23–399.21) |
| **MIG** | Monokine Induced by Gamma Interferon | Chemokine | Immune cell recruitment | pg/ml | 86.12 (61.69–120.99) |
| **MIP-1α** | Macrophage Inflammatory Protein-1 Alpha | Chemokine | Immune cell recruitment | pg/ml | 15.47 (12.38–17.94) |
| **MIP-1β** | Macrophage Inflammatory Protein-1 Beta | Chemokine | Immune cell recruitment | pg/ml | 69.56 (53.09–88.50) |
| **PAI-1** | Plasminogen Activator Inhibitor-1 | Protein | Clot regulation | ng/ml | 1.82 (1.18–3.49) |
| **SAA** | Serum Amyloid A | Protein | Inflammation | ng/ml | 4075.30 (2542.36–7173.55) |
| **sCD30** | Soluble Cluster of Differentiation 30 | Protein | Immune activation | pg/ml | 514.19 (79.00–1426.57) |
| **sVCAM-1** | Soluble Vascular Cell Adhesion Molecule-1 | Protein | Immune cell adhesion | ng/ml | 523.03 (446.99–606.93) |
| **TGF-α** | Transforming Growth Factor Alpha | Protein | Cell growth | pg/ml | 1.90 (1.53–2.48) |
| **TGF-β** | Transforming Growth Factor Beta | Cytokine | Immune regulation | pg/ml | 2654.59 (1468.95–4153.87) |
| **TNFR1** | Tumor Necrosis Factor Receptor I | Protein | Inflammatory signaling | pg/ml | 3473.04 (2924.55–4066.26) |
| **TNF-α** | Tumor Necrosis Factor Alpha | Cytokine | Inflammation | pg/ml | 1.21 (0.99–1.58) |
| **TRAIL** | TNF-Related Apoptosis-Inducing Ligand | Cytokine | Cell death | pg/ml | 120.04 (102.55–150.23) |

\*IQR = interquartile range; \*\*number of participants CMV IgG-, sample prevalence in parentheses

#### Biomarker Processing

Plasma biomarkers were quantified by immunoassay, and custom multiplex assays were designed to optimize sample use. Ten assay kits were used to analyze the 48 candidate biomarkers; see **Supplemental Table 2** for assay descriptions and precision. A Human Control Plasma (HCP) sample, pooled from 10 healthy subjects, was run in duplicate on each plate to assess inter-assay variability of each assay. The mean of the control samples for all assays – plus or minus two standard deviations (SD) – was defined as the acceptable control range. Concentrations below the lower level of detection (LLOD) were imputed as half the LLOD.

Except for CMV IgG, all biomarker measures were continuous. The semi-quantitative plasma CMV IgG biomarker was reported either as a numeric concentration (if >10 EU/ml) or as “Negative.” To incorporate this variable into the analysis, we generated two derived variables: (1) a numeric variable that retained the reported concentration, and if “Negative,” was set to 0; and (2) a binary indicator equal to 1 for “Negative” and 0 otherwise. This approach preserved quantitative information while capturing the distinction between detectable and undetectable values, allowing the model to select the most informative representation. Overall, this resulted in 49 biomarker-related variables. For subsequent LASSO selection, all continuous concentrations were natural log-transformed and then z-score normalized.

### 2.3 Pain Outcomes

Pain intensity was assessed pre-operatively and 6 months post-operatively using the PROMIS Pain Intensity Scale v1.0 3s, which includes three items evaluating pain intensity experienced over the prior seven days. Pain interference was assessed at the same time points using the PROMIS Pain Interference Scale – Short Form 6A, which includes six items assessing the extent to which pain interfered with daily activities over the prior seven days. Both scales use a 5-point Likert response format, ranging from 0 (‘no pain’ or ‘not at all’) to 4 (‘very severe’ or ‘very much’). Final scores for both outcomes were calculated using the HealthMeasures Scoring Service, powered by Assessment Center^SM^.

### 2.4 Statistical Analysis

All statistical analyses were run using R Version 4.4.0 [97].

#### Expected Recovery Differential (ERD) Derivation

ERDs are derived by comparing each participant’s observed recovery to the expected recovery predicted by a model that accounts for demographic, comorbidity, reserve, and condition-related factors [17]. In other words, the ERD score reflects how much better or worse a given patient’s CPSP was observed to be, compared to what clinicians might have expected. ERD derivation inputs and processes are illustrated in **Figure 1** and listed in **Supplemental Table 1**. A total of 29 pre-operative variables were used to derive pain intensity and interference ERDs, and included the following: baseline pain (intensity or interference), demographic factors (e.g., age, sex), health factors (e.g., BMI, comorbid conditions), physical performance (e.g., distance walked, grip strength), cognitive performance (e.g., processing speed, executive function), psychosocial factors (e.g., depressive symptoms, emotional support), and surgical factors (e.g., blood loss, anesthesia type/time).

**Figure 1.**
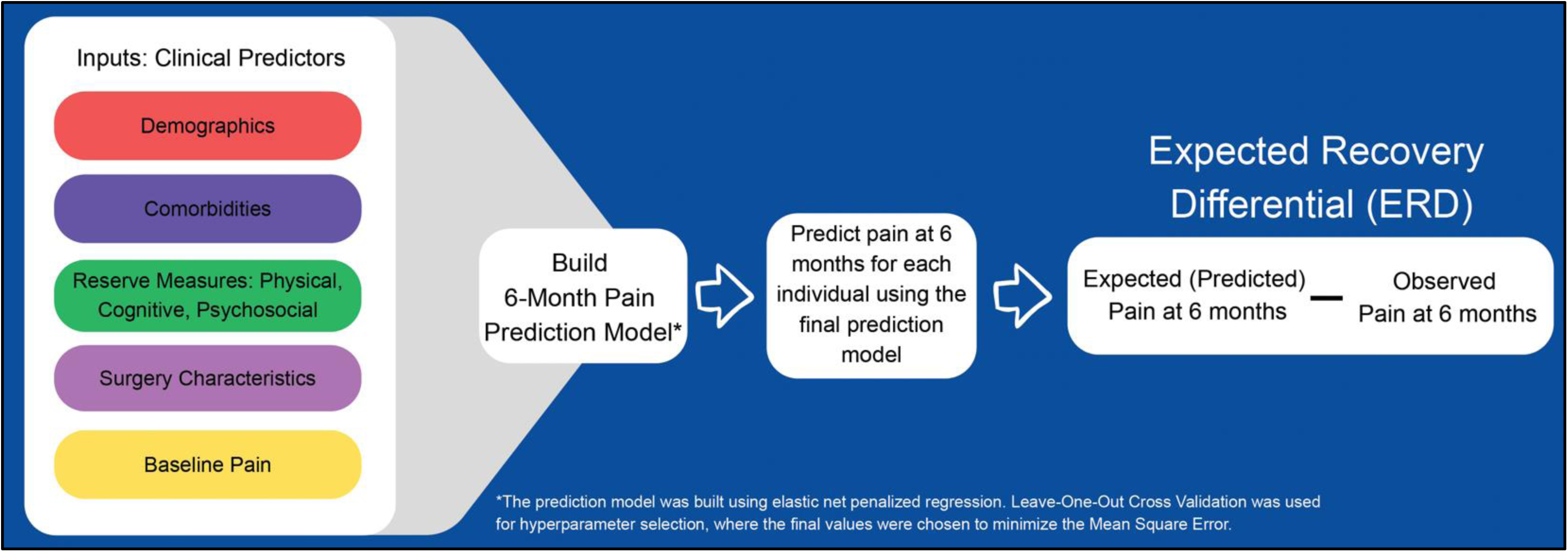
ERD Derivation Diagram. Clinical predictors are further described in Supplemental Table 1.

Elastic net penalized regression including the covariates listed above was used to create prediction models for 6-month pain intensity and pain interference. For each model, predictors with zero variation in the sample (i.e., all 0’s or all 1’s for a binary predictor) were removed, and all continuous variables were centered/scaled. To determine the optimal penalty parameter (λ) and elastic-net mixing parameter (α), Leave One Out Cross Validation was used, and the combination of parameters with the lowest mean squared error was chosen for the final model. Note that setting α =1 results in a LASSO regression, whereas setting λ = 0 results in a ridge regression [140]. Values of λ ≥ 0 and values of α in [0,1] were considered. The coefficients resulting from these final models were used to predict the pain outcome for each participant at 6 months based on their individual characteristics. The ERD was then calculated by subtracting this predicted pain outcome at 6 months from the observed pain outcome at 6 months (**Figure 1**).

For ease of interpretation, the final direction of the ERDs was changed such that a higher ERD indicates greater improvement in pain than expected (i.e. better pain resilience), and a lower ERD indicates worse improvement in pain than expected (i.e. worse pain resilience). Additionally, all ERD values were centered and scaled to standardize the distributions. For each outcome, only those with available 6-month values were included in the analyses (**Figure 2**); pain intensity ERD was computed for 169 participants, pain interference ERD for 170 participants.

**Figure 2.**
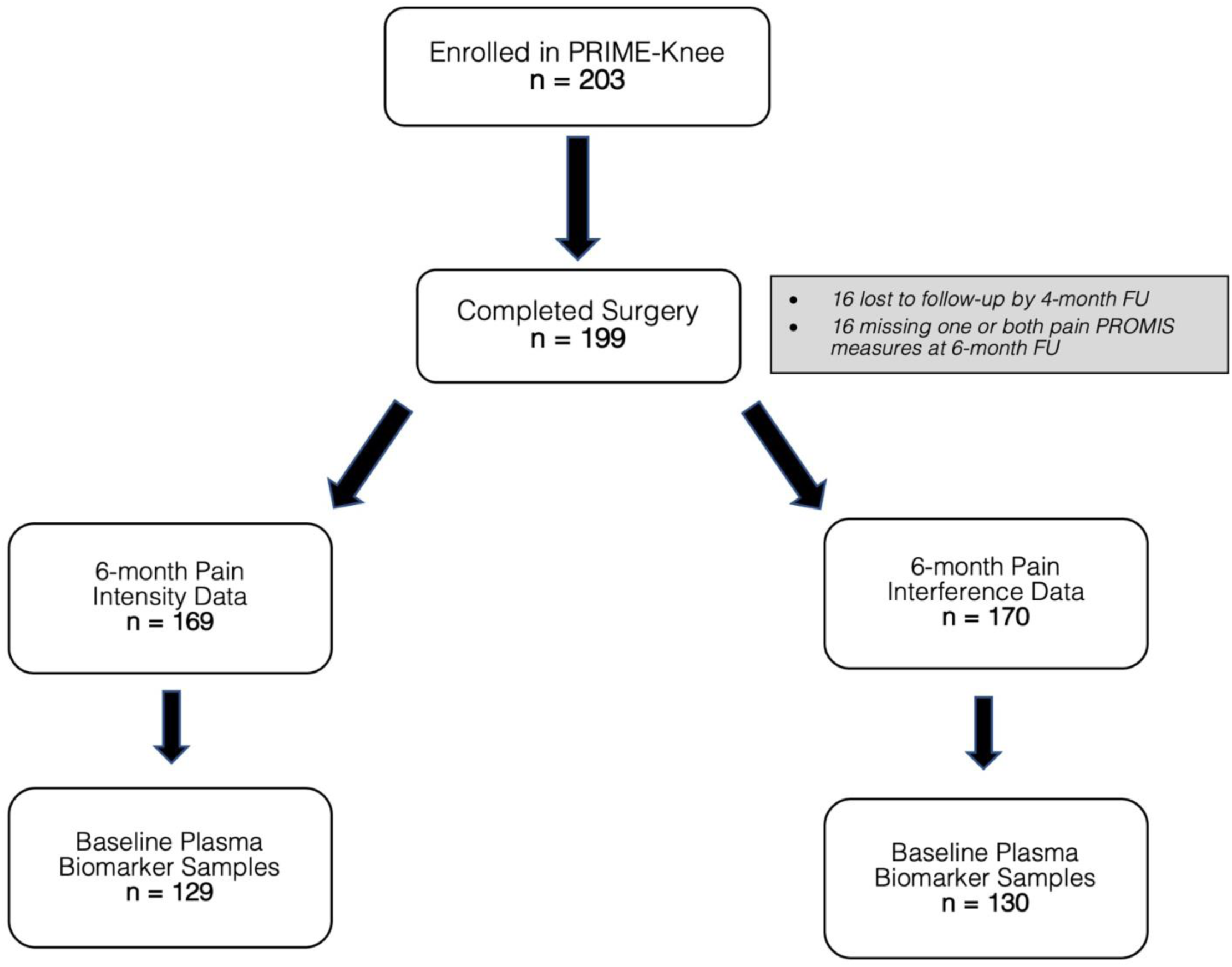
CONSORT Diagram.

#### Predictive Biomarker Discovery

The following exploratory stability selection procedure was used to select a stable set of biomarkers associated with pain intensity and pain interference ERDs. First, we resampled the data 200 times by bootstrapping (sampling with replacement) and 200 times using a complementary pairs method (random half-splitting) [108]. On each resampled dataset, we ran five LASSO-penalized regression models with different penalty parameters. These penalty parameters were calibrated using a full-dataset reference model resulting in exactly 5,10, 15, 20, and 25 plasma biomarkers having a non-zero coefficient, and thus approximately those sparsity levels across resamples when modeling the ERD outcome. This resulted in ten different modeling configurations, defined by the two resampling methods and the five target sparsity levels (i.e., desired numbers of non-zero coefficients) used in the LASSO models. For each of these ten modeling configurations, we calculated the selection probability (times out of 200) of each biomarker being selected as a non-zero parameter estimate. The biomarkers with the top 10 selection probabilities from each modeling configuration were considered in the interim stable set and used for further analysis. The union of all ten interim stable sets (i.e., one for each configuration) resulted in one grand stable set for each ERD outcome. The associated stability score represents the number out of 10 stable sets that each biomarker was selected.

Forest plots were constructed to illustrate the adjusted effect estimates for each candidate biomarker. Adjustment included all biomarkers in the grand stable set plus a post-operative event adjustment variable. Specifically, patients were asked the following question at each post-operative check in: “Have you had an illness/injury that has prevented you from performing your regular activities for one day or more?” This event variable was considered affirmative if patients provided at least one yes at 1, 2, 4 or 6 months. Since the goal of this analysis was hypothesis generation, and we used a data-driven selection process that yields naïve variance estimators from linear regression, typical post-selection procedures are invalid and thus no quantitative measure of uncertainty was reported. Participants were only included in this biomarker analysis if they had both a calculated ERD and a baseline plasma sample available. This resulted in 129 participants for the pain intensity ERD analysis and 130 for the pain interference ERD analysis (**Figure 2**).

#### Protein–Protein Interaction Analysis (PPI)

Stably-selected protein biomarkers with the highest stability score (10 selections out of 10 possible) for each ERD domain were entered into the STRING PPI Network (version 12.0; https://string-db.org/) PPI network. A third network combined biomarkers from both ERD domains to examine cross-connectivity independent of prediction. Biomarkers were entered into a Homo sapiens model using the full STRING network with a minimum medium confidence interaction score (≥0.400). Number of edges were compared against expected edges per random selection; which reflects the mean number of edges expected for the same number of proteins drawn at random from the full human proteome. Planned network measures included the number of nodes (proteins) and edges (interactions), edge confidence (≥0.400 = medium, ≥0.700 = higher, ≥0.900 = highest), average node degree (individual interactions per biomarker), and PPI enrichment p-value (probability nodes in network are not random and edges are significant) [116].

## 3. RESULTS

The average age of the participants was 71 years; the majority identified as female (60%) and white (82.9%). Additional sample information is available in **Table 2**.

**Table 2.** Patient Descriptive Statistics.

|  |  |  |  |
| --- | --- | --- | --- |
| **Age at Surgery Date (years)** |  | **Opiate** |  |
| Mean (SD) | 71.4 (6.20) | Yes | 20 (11.8%) |
| Median [Min, Max] | 70.7 [60.1, 84.8] | No | 145 (85.3%) |
| **Race** |  | Missing | 5 (2.9%) |
| White | 141 (82.9%) | **NSAID** |  |
| Black or African American | 22 (12.9%) | Yes | 67 (39.4%) |
| Asian | 2 (1.2%) | No | 98 (57.6%) |
| Native American or Alaska Native | 0 (0%) | Missing | 5 (2.9%) |
| Native Hawaiian or Pacific Islander | 0 (0%) | **3MS** |  |
| More than one race | 4 (2.4%) | Mean (SD) | 93.1 (6.49) |
| Unknown or Not Reported | 1 (0.6%) | Median [Min, Max] | 95.0 [62.0, 100] |
| **Ethnicity** |  | Missing | 4 (2.4%) |
| Not Hispanic or Latino | 161 (94.7%) | **15 Item word List** |  |
| Hispanic or Latino | 7 (4.1%) | Mean (SD) | 6.47 (1.93) |
| Unknown or Not Reported | 2 (1.2%) | Median [Min, Max] | 6.00 [2.00, 11.0] |
| **Sex** |  | Missing | 1 (0.6%) |
| Female | 102 (60.0%) | **Trail Making Test A (sec)** |  |
| Male | 68 (40.0%) | Mean (SD) | 36.5 (18.5) |
| **Education Level (years)** |  | Median [Min, Max] | 33.0 [16.0, 180] |
| Mean (SD) | 16.2 (2.36) | Missing | 1 (0.6%) |
| Median [Min, Max] | 16.0 [10.0, 20.0] | **Trail Making Test B (sec)** |  |
| Missing | 1 (0.6%) | Mean (SD) | 91.1 (44.7) |
| **BMI** |  | Median [Min, Max] | 79.0 [37.5, 300] |
| Mean (SD) | 32.0 (5.34) | Missing | 6 (3.5%) |
| Median [Min, Max] | 32.4 [19.9, 42.6] | **Gait Speed (m/sec)** |  |
| **Vascular Disease** |  | Mean (SD) | 1.00 (0.256) |
| Yes | 44 (25.9%) | Median [Min, Max] | 1.05 [0.349, 1.54] |
| No | 121 (71.2%) | **3-minute Walk (ft)** |  |
| Missing | 5 (2.9%) | Mean (SD) | 604 (178) |
| **Chronic Liver Disease** |  | Median [Min, Max] | 633 [0, 1210] |
| Yes | 5 (2.9%) | **Grip Strength (kg)** |  |
| No | 159 (93.5%) | Mean (SD) | 29.2 (9.98) |
| Missing | 6 (3.5%) | Median [Min, Max] | 28.0 [2.00, 60.0] |
| **Heart Disease** |  | **PHQ-9** |  |
| Yes | 9 (5.3%) | Mean (SD) | 3.25 (3.66) |
| No | 156 (91.8%) | Median [Min, Max] | 2.00 [0, 24.0] |
| Missing | 5 (2.9%) | Missing | 16 (9.4%) |
| **Diabetes** |  | **Resilience Scale** |  |
| Yes | 35 (20.6%) | Mean (SD) | 150 (25.0) |
| No | 130 (76.5%) | Median [Min, Max] | 154 [25.0, 175] |
| Missing | 5 (2.9%) | Missing | 35 (20.6%) |
| **History of Treated Non-Skin Cancer** |  | **Post-Operative Illness/Injury\*** |  |
| Yes | 11 (6.5%) | Yes | 56 (32.9%) |
| No | 153 (90.0%) | No | 114 (67.1%) |
| Missing | 6 (3.5%) |  |  |
| **Antidepressant** |  |  |  |
| Yes | 34 (20.0%) |  |  |
| No | 131 (77.1%) |  |  |
| Missing | 5 (2.9%) |  |  |

### 3.1 Biomarker Selection

Biomarker abbreviations, names, descriptions, and baseline values are listed in **Table 1**. Biomarker selection was performed for each patient with both an ERD outcome and plasma samples (129 of 169 participants for pain intensity ERD; 130 of 170 participants for pain interference ERD; see **Figure 2**). The exploratory stability selection framework selected the most stable presurgical biomarkers associated with each ERD. Two biomarkers met the highest selection criteria (selected 10 /10 times in interim stable sets) for both pain intensity and pain interference ERDs: IL-5 (positively associated) and LBP (negatively associated).

In addition to IL-5 and LBP, four other biomarkers met the highest selection criteria and were negatively associated with pain intensity ERD (**Figure 3a**): CMV IgG-, MIP-1β, IL-12p70, and sCD30. Two biomarkers met the highest selection criteria and were positively associated with the pain intensity ERD: IFNα-2a and LIF. In addition to IL-5 and LBP, four biomarkers met the highest selection criteria and were negatively associated with pain interference ERD (**Figure 3a**): LPS, activin A, IL-8, and SAA. One biomarker, IL-7, met the highest selection criteria and was positively associated with the pain interference ERD.

**Figure 3.**
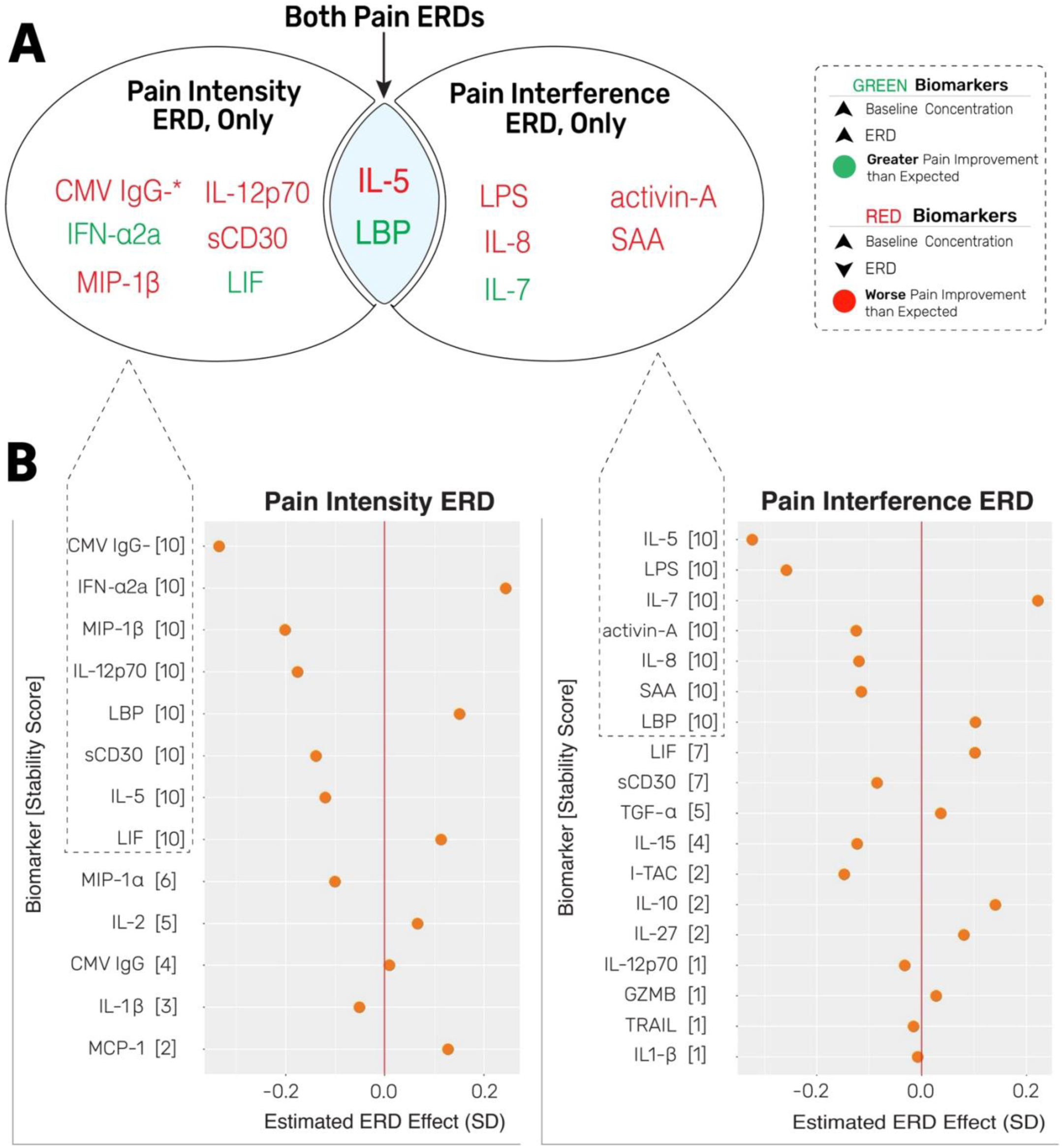
Most Stably Selected Biomarkers by Pain ERD. A) Unique versus shared stable biomarker candidates by pain ERD; restricted to biomarkers with highest stability score (selected 10/10 times) via exploratory stability selection. B) Biomarker regression estimates by pain ERD, after adjusting for all candidate biomarkers in the grand stable set plus post-operative illness/injury variable (see Table 2). Scaled units are standard deviation (SD);Values indicate increase or decrease in ERD relative to average outcome, for every 1 standard deviation increase in ln biomarker concentration. * = CMV IgG-is a binary variable, i.e. testing negative for CMV IgG (yes/no) was associated with 0.3 SD lower ERD (worse pain improvement than expected) compared to testing positive for CMV IgG).

### 3.2 Biomarker Effect Size Estimates

**Figure 3b** provides forest plots for all biomarkers present in the grand stable set for each pain ERD; effect size estimates reflect the adjustment for all other biomarkers plus the post-operative event variable. CMV IgG– was the stable biomarker with the greatest effect size and negative association with pain intensity ERD; as a binary variable, this means that CMV IgG– = ‘yes’ (i.e. testing negative for CMV IgG) was associated with 0.33 SD lower ERD (i.e. worse than expected postsurgical pain intensity improvement) compared to CMV IgG– = ‘no’ (testing positive for CMV IgG). MIP-1β demonstrated a similar but weaker negative association with pain intensity ERD; i.e. a 1 SD increase in its baseline ln concentration was associated with 0.20 SD lower (worse) ERD. IFNα-2a was the stable biomarker with the strongest positive association with pain intensity ERD; a 1 SD increase in its baseline ln concentration was associated with 0.24 SD higher ERD (i.e. greater than expected postsurgical pain intensity improvement).

IL-5 was the stable biomarker with the strongest negative association with pain interference ERD; a 1 SD increase in its baseline ln concentration was associated with 0.32 SD lower ERD (i.e. worse than expected postsurgical pain interference improvement). LPS also met the highest selection criteria but with slightly weaker prediction; a 1 SD increase in baseline LPS ln concentration was associated with 0.26 SD lower ERD. IL-7 was the stable biomarker with the strongest positive association with pain interference ERD; a 1 SD increase in its baseline ln concentration was associated with 0.22 SD higher ERD (i.e. greater than expected postsurgical pain interference improvement).

### 3.3 Protein-protein interaction analysis

Protein-protein interaction analysis (PPI) was performed to identify physical contacts and functional associations between proteins associated with pain intensity and pain interference, to gain a deeper understanding of the biological processes involved. Biomarkers with the highest selection criteria were considered for PPI. For pain intensity ERD **(Figure 4A)**, preliminary nodes were CMV IgG-, MIP-1β, IL-12p70, sCD30, IFN-α2a, LIF, IL-5, and LBP. While it met the selection criteria, CMV IgG-was excluded as it is a clinical test indicating the absence of antibody-mediated immune response rather than a single gene-encoded protein. Similarly, IL-12p70 is a protein heterodimer and thus requires representation by its P35 (IL-12A) and P40 (IL-12B) gene-encoded subunits [123]. All nodes demonstrated at least one known interaction, and 13 total interactions were found, substantially higher connectivity than expected (random selection = 1 interaction). MIP-1β had the highest connectivity at 6 edges. Average node degree was 3.25 (>3 interactions per biomarker). Independent of IL-12A and IL-12B subunits that together form the IL-12p70 heterodimer, the strongest edge confidence was between MIP-1β and IL5 (0.826 or exceeding ‘high’ confidence). PPI enrichment p-value was p < 1.0 × 10⁻^13^ (extremely low likelihood of network connectivity by chance).

**Figure 4.**
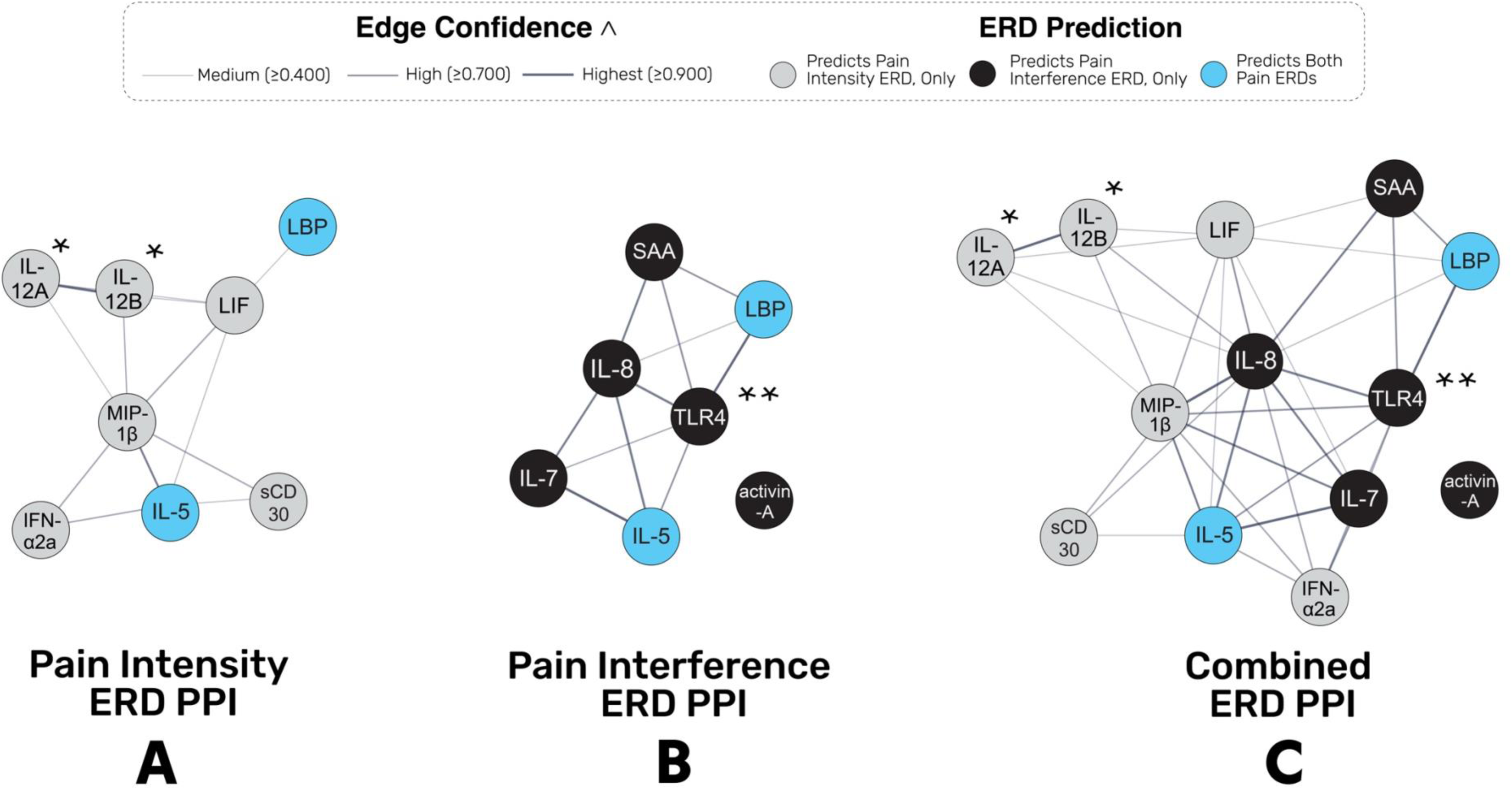
Protein-protein interaction analysis (PPI) of most stably selected biomarkers by pain intensity ERD (A), pain interference ERD (B), and combined ERD (C) networks. Restricted to biomarkers with highest selection criteria (selected 10/10 times);* Subunits of the non-gene coded heterodimer, IL12p70; ** downstream canonical receptor of the non-gene coded glycolipid, LPS; ^ confidence score based on the approximate probability of a predicted link between proteins in the same metabolic map, in the bioinformatics (KEGG) database [116].

For the pain interference ERD PPI (**Figure 4B**), preliminary nodes were LPS, LBP, activin A, IL-8, SAA, IL-7, and IL-5. LPS (non-protein glycolipid) was represented by its downstream canonical receptor, TLR4 [92]. Except for activin A, all nodes demonstrated at least one known interaction. A total of 11 interactions were found, which was substantially higher connectivity than expected (random selection = 2 interactions). IL-8 and TLR4 accounted for the most interactions at 5 apiece. Average node degree was 3.14 (>3 interactions per biomarker). Independent of TLR4 and LBP (a strong LPS-TLR4 mediator [92]), the ‘highest’ edge confidences were observed between IL-5 and IL-7 (0.995), IL-8 and TLR4 (0.933), and IL-5 and IL-8 (0.932). PPI enrichment p-value was p < 1.0 × 10⁻^5^ (low likelihood of network connectivity beyond chance).

For the combined ERD PPI (**Figure 4C)**, all nodes listed above were entered into one network; except for activin A, all nodes demonstrated at least one known interaction. Twelve interactions added to the 24 already found in separate ERD models, for a total of 36 interactions or substantially higher connectivity than expected (random selection=4 interactions). IL-8 demonstrated the highest connectivity at 11 edges, followed by MIP-1β with 9 edges. Average node degree was 5.54 (>5 interactions per biomarker). Strong cross-connectivity (between unique ERD predictors) was observed for IL-8 and MIP-1β (0.988 or exceeding ‘highest’ confidence), and IL-8 and LIF (0.731 or exceeding ‘high’ confidence). PPI enrichment p-value was p < 1.0 × 10⁻^16^ (extremely low likelihood of network connectivity by chance).

### 3.4 Post-hoc analyses

#### Post-hoc sensitivity analyses

A post-hoc analysis was performed to determine the extent to which pre-operative opioid or NSAID use modified associations between biomarkers and pain ERDs. We performed linear regression modeling to test for interactions and found no statistically significant evidence (at *α*=0.05) of varied effect of biomarker on pain ERDs for different medication use categories.

#### Post-hoc PPI

Unanticipated findings of CMV negative (IgG-) status as a predictor of worse pain intensity ERD coincided with a recently published seminal study, whereby CMV seropositivity was protective for mortality and cytotoxicity among patients receiving immunotherapy for malignant melanoma [78]. The study proposed TBX21 (encoding T-bet) as the primary transcription factor explaining CMV seropositivity protective effects; this finding was corroborated by previous data on signaling pathways for TBX21 and CD8+ T cell prevalence and ratios in persons with and without CMV [43,100]. Accordingly, we supplemented the combined ERD PPI network with TBX21, which is gene-encoded and available in STRING databases (**Figure 5A**). TBX21 demonstrated five edges, suggesting known interactions with IL-5, IL-7, IL-8, MIP-1β, and TLR4. Further, a ‘high’ edge confidence was observed between TBX21 and IL-5 (0.839). PPI enrichment p-value was p < 1.0 × 10⁻^16^, suggesting an extremely low likelihood of network connectivity by chance.

**Figure 5.**
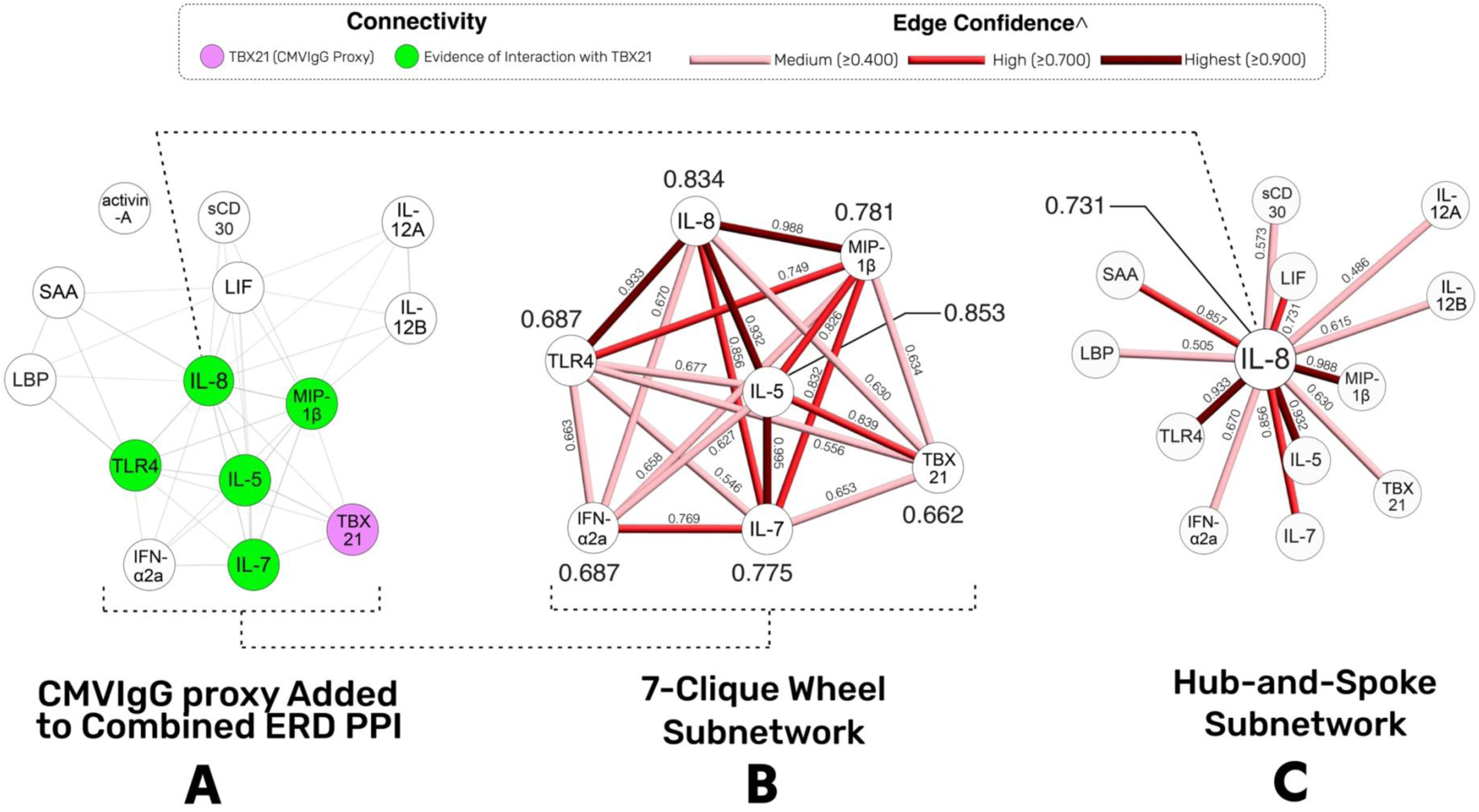
Post-hoc protein-protein interaction analysis (PPI; A), and subnetwork extraction (B,C), to visualize and quantify connectivity. Numbers adjacent to edges indicate single edge confidence score; italicized numbers indicate average edge confidence score by node; ^ color corresponding to edge confidence score, based on the approximate probability of a predicted link between proteins in the same metabolic map, in the bioinformatics (KEGG) database [116].

### 3.5 Subnetwork Extraction

A coherent five-clique wheel graph subnetwork was observed in the combined ERD PPI (**Figure 4C**); adding TBX21 (CMV IgG-proxy) further enhanced the subnetwork symmetry to form a 7-clique wheel graph (**Figure 5A**). Subnetworks potentially highlight shared biological function among proteins; topological examination of these subnetworks may help guide biomarker exploration [16,86,125]. To this end, a simple subnetwork extraction was performed to visualize and quantify the connectivity of the 7-clique wheel graph formed by IL-5, IL-7, IL-8, MIP-1β, IFN-α2a, TLR4 (LPS proxy), and TBX21 (**Figure 5B**). Average edge confidence for the subnetwork was 0.752 or ‘high.’ Eight of 20 nodes were categorized as ‘high’ or ‘highest’ edge confidence. Specific to each node, four had a ‘high’ average edge confidence in the following rank-order: IL-5 (hub), IL-8, MIP-1β, and IL-7.

Lastly, IL-8 was found to have 12 edges, suggesting an interaction with nearly every stably-selected biomarker meeting 10/10 selection criteria. A simple subnetwork extraction was performed to visualize degree centrality and connectivity (**Figure 5C**). The average edge confidence for IL-8 was 0.731, classified as ‘high.’ Overall, IL-8 showed a relatively high degree of centrality compared to other nodes, with a weighted hub-and-spoke subnetwork in which 6 of 12 edges were determined to have ‘high’ or ‘highest’ confidence. IL-8 also demonstrated ‘high’ edge confidence with SAA and LIF, in addition to edges identified in the wheel-graph subnetwork of IL-5, IL-7, TL4, and MIP-1β.

## 4. DISCUSSION

In a prospective, longitudinal cohort of patients undergoing TKA (PRIME-Knee [19,127]), we identified 13 presurgical immune biomarkers that met the highest selection criteria (selected in 10/10 interim stable sets) for having associations with 6-month postsurgical pain recovery ERDs. Two biomarkers – LBP and IL-5 – were associated with both pain intensity and pain interference ERDs. With the exception of activin A, the most stably selected biomarker group demonstrated high network connectivity. Two highly symmetrical subnetworks centered on IL-5 and IL-8. Collectively, these results introduce a preliminary set of highly interactive (biologically plausible) biomarkers worthy of scrutiny in future CPSP biosignature derivation studies. Notable findings are discussed below, with the caveat that all mechanistic reasoning remains speculative until tested further.

### 4.1 Preliminary peripheral biosignature of CPSP resilience

A high proportion of the most stably-selected biomarkers operate within or downstream of the LPS-TLR4 signaling pathway, which is the body’s primary defense against Gram-negative bacterial infections [31,137]. Accumulating evidence also implicates LPS-TLR4 signaling in a shortened healthspan starting with gut dysbiosis, which facilitates a systemic rise in LPS (endotoxemia) and initiates low-grade age-related inflammation and cardiometabolic disease [13,73,79,101]. Dysbiosis-endotoxemia is also a known culprit in arthritis-related joint disease [12,49,50,66]. Members of our team conceptualized this OA disease pathway to be a ‘two-hit’ model: 1) systemic LPS facilitates TLR4 → NF-κB → inflammatory signaling (peripheral hyperalgesia); 2) knee joint damage disease-associated molecular patterns (DAMPs) facilitating TLR4 over-activity and hyper-inflammation (higher/persistent peripheral hyperalgesia) [48,50,126].

LBP sits at the apex of the LPS-TLR4 pathway, shuttling LPS to TLR4 receptors [93]. Higher LBP levels are linked to OA progression [49,50,129] and small fiber neuropathy (SFN) [56,115], consistent with the two-hit model. In this study, however, presurgical LBP was *inversely* associated with CPSP, a relationship consistent with an OA disease-*resolution* framework rather than one of disease progression. TKA removes OA-related joint damage and the associated ‘second-hit’ DAMPs [48]. Although DAMPs can re-emerge after acute surgical trauma, or later implant wear, neither is likely at 6 months [72,90,117,119]. With OA-related DAMPs eliminated, TLR4 overactivity should decline, reducing the hyper-inflammatory state that contributed to presurgical chronic knee pain.

Taken together, these findings suggest that LBP may serve as a proxy ‘canary in the coal mine’ for TKA responsivity and CPSP resilience. Because the LPS-TLR4 pathway drives peripheral hyperalgesia, it *should* improve when TKA removes joint-derived DAMPs [48]). Consistent with this, higher LBP was also associated with better-than-expected pain interference outcomes. When joint-loading movement is perceived as ‘less painful’ after TKA, this often — though not always [82] — indicates successful elimination of the peripheral pain generator (TKA ‘hit the mark’) [9,11,23,76,99]. The previously reported association between LBP and SFN [56] further strengthens this biosignature. SFN is a peripherally dominant condition in which symptom severity tracks relatively linearly with tissue pathology (e.g., intra-epidermal nerve fiber loss) [6,24,25,36,84,115,121].

Other LPS-TLR4 biomarkers, LPS, IL8, IL12p80, and SAA, were associated with *worse*-than-expected postsurgical pain recovery. This is biologically plausible given the high pleiotropy of the LPS-TLR4 system. LBP illustrates this well: when LPS is low and then acutely spikes, LBP facilitates TLR4 signaling by transferring LPS downstream [122]. However, when LPS remains chronically high, LBP instead inhibits LPS transfer to preserve TLR4 sensitivity [41]. A second consideration is the ubiquity of the LPS-TLR4 pathway. Distinct tissue- and organ-specific variants exist across both peripheral and central systems. Consequently, circulating LPS-TLR4 biomarkers can reflect *both* peripheral and central pain mechanisms, which may explain why some markers align with poorer postsurgical pain trajectories.

### 4.2 Preliminary central biosignature of CPSP susceptibility

Knee-derived DAMPs activate TLR4 receptors both locally and within the dorsal root ganglion (DRG). [3,10,15,65]. When this afferent barrage is sufficiently intense or prolonged, DRG-resident DAMPs progressively sensitize local ion channels and initiate neuropathic pain pathogenesis [2,10,42,63,71,120,130]. Temporal dynamics are critical: spinal TLR4 signaling activates microglia during the acute phase and astrocytes in the chronic phase [3,12,18,72]. If afferent input resolves early, neuropathic pain is less likely to develop; however, once firing becomes persistent, neuropathic pain becomes less modifiable and may outlast the resolution of peripheral hyperalgesia [15,63,120]. A rodent surgical incision/retraction model demonstrates a similar cell-specific window to chronic CPSP development driven that by a central TLR4 → DAMPs pathway [42].

Additional findings reinforce this central mechanism. IL-8 was associated with worse-than-expected postsurgical pain interference ERD and demonstrated high connectivity and centrality in network analyses. IL-8 (CXCL8) is abundant in synovial fluid and linked to joint destruction [34,80]. Neutrophils—key drivers of hyper-inflammation and joint damage—are strongly recruited by IL-8, the most potent neutrophil-attractant chemokine in humans [14]. Unlike most cytokines, IL-8 signals through G-Protein-coupled receptors (e.g., CXCR1/CXCR2) [85,109], which, like TLR4, are expressed peripherally and centrally on immune cells and peripheral sensory neurons, including the DRG. A growing body of evidence links IL-8 to neuropathic pain and centrally mediated autoimmune diseases [14]. MIP-1β, although less directly tied to TLR4 → DAMPs, was also associated with pain ERDs and has been previously linked to neuropathic and visceral pain [52,103,104].

Finally, accumulating evidence points to neutrophils as key mediators of the acute-to-chronic pain transition and OA progression [47,91]. Together, these findings support the biological plausibility of a centrally dominant IL-8-based biosignature of CPSP susceptibility, consistent with the growing recognition of neuropathic pain prevalence in knee OA and CPSP [24,105,107]. Because neuropathic pain is less modifiable and less responsive to TKA than peripheral hyperalgesia [57,131], the association between higher presurgical IL-8 and worse pain inference ERD is mechanistically aligned.

### 4.3 Preliminary indication of adaptive immunity dysregulation

The two biosignatures described above primarily reflect innate immune processes, which have a well-established role in pain. However, two biomarkers in this study implicate adaptive immunity, which operates more slowly but with greater antigen specificity and involves lymphocyte-mediated memory and effector signaling. IL-5, a canonical TH2 cytokine associated with eosinophilic inflammation in upper airway and dermatologic disorders [118], was the only biomarker besides LBP associated with both ERDs, and in a worse-than-expected direction. IL-5 also emerged as the central ‘hub’ of the highly connected 7-clique wheel subnetwork. Prior work shows women with fibromyalgia exhibit higher monocyte-stimulated IL-5, whereas animal studies found IL-5 to be protective [59,77]. Given the pleiotropic nature of immune pathways and the chronic pain status of this cohort, alignment with fibromyalgia findings is biologically plausible.

The second biomarker, CMV seronegativity status, demonstrated the strongest association with worse-than-expected pain intensity ERD. This was an unanticipated finding, because CMV IgG seronegativity is traditionally considered protective against cardiovascular disease and mortality [89]. Emerging evidence complicates this narrative [37,83,138]. For example, CMV *seropositivity* may protect against multiple sclerosis in patients with Epstein-Barr Virus (EBV) [40]. Among individuals with malignant melanoma, CMV seropositivity is associated with lower long-term mortality nearly a decade later and a more resilient pre-treatment immune profile during anti-PD-1 therapy [78].

In CPSP, adaptive immunity findings must be interpreted cautiously due to limited literature and the exploratory nature of this study. Still, adaptive immunity involves competing TH1 and TH2 functions. T Helper Cells 1 (T_H_1) mediate intracellular defense, whereas Helper T Cells 2 (T_H_2) cells coordinate antibody-mediated responses to extracellular threats [139]. CMV seropositivity increases T_H_1 memory inflation and cytotoxic effector function, maintaining heightened vigilance. Recent findings linking CMV seropositivity to protection against melanoma and EBV support this interpretation [40,78]. In parallel, IL-5 reflects T_H_2 activity—allergic and atopic reactions against pathogens. The current pattern—higher IL-5 and CMV seronegativity predicting worse outcomes—suggests a shift toward a T_H_2-dominanat endotype, potentially contributing to CPSP susceptibility.

### 4.4 Strengths and Limitations

Several limitations should be considered. Post-operative medication use was not examined, although pre-operative use — an established predictor — did not moderate findings. Biomarker analyses were limited to plasma, representing only a fraction of the immune processes relevant to CPSP. Comparisons with studies using different assay platforms (e.g., proteomics) are challenging [39]. Detailed post-operative rehabilitation data were not collected, although physical characteristics were included in resilience models and surgical/clinical practices were relatively homogeneous. Follow-up ended at 6 months, limiting long-term inference; however, both our data and prior cohorts indicate pain outcomes typically plateau by this time.

This study also has notable strengths. We evaluated a broad panel of presurgical aging, OA, and pain biomarkers; assessed post-operative pain through both susceptibility and resilience frameworks; used rigorous quantitative post-operative measures such as ERD; quantified both pain intensity and interference outcomes; applied a robust stability-selection framework to identify biomarker associations with CPSP; and included PPI and subnetwork extraction to interrogate biomarker connectivity.

### 4.5 Conclusions

A highly stable set of presurgical biomarkers was associated with pain recovery after TKA. The direction and magnitude of these associations, combined with the high connectivity and symmetry of biomarker networks, support the biological plausibility of preliminary CPSP biosignatures of susceptibility and resilience. Future basic and clinical studies are needed to derive and validate these signatures for clinically meaningful risk stratification or treatment tailoring. Until then, findings remain exploratory and are intended to generate hypotheses and stimulate field dialogue.

## Data Availability

All data produced in the present study are available upon reasonable request to the authors.

## Conflict of Interest Statement

The authors have no conflicts of interest to declare.

## Acknowledgements

The study was funded by NIH National Institute on Aging (NIA) under Award Numbers UH3AG056925, P30AG072958, P30AG028716, and K76AG074943. Deidentified individual participant data will be made available upon reasonable request to the corresponding author.

---

**Supplemental Table 1.**
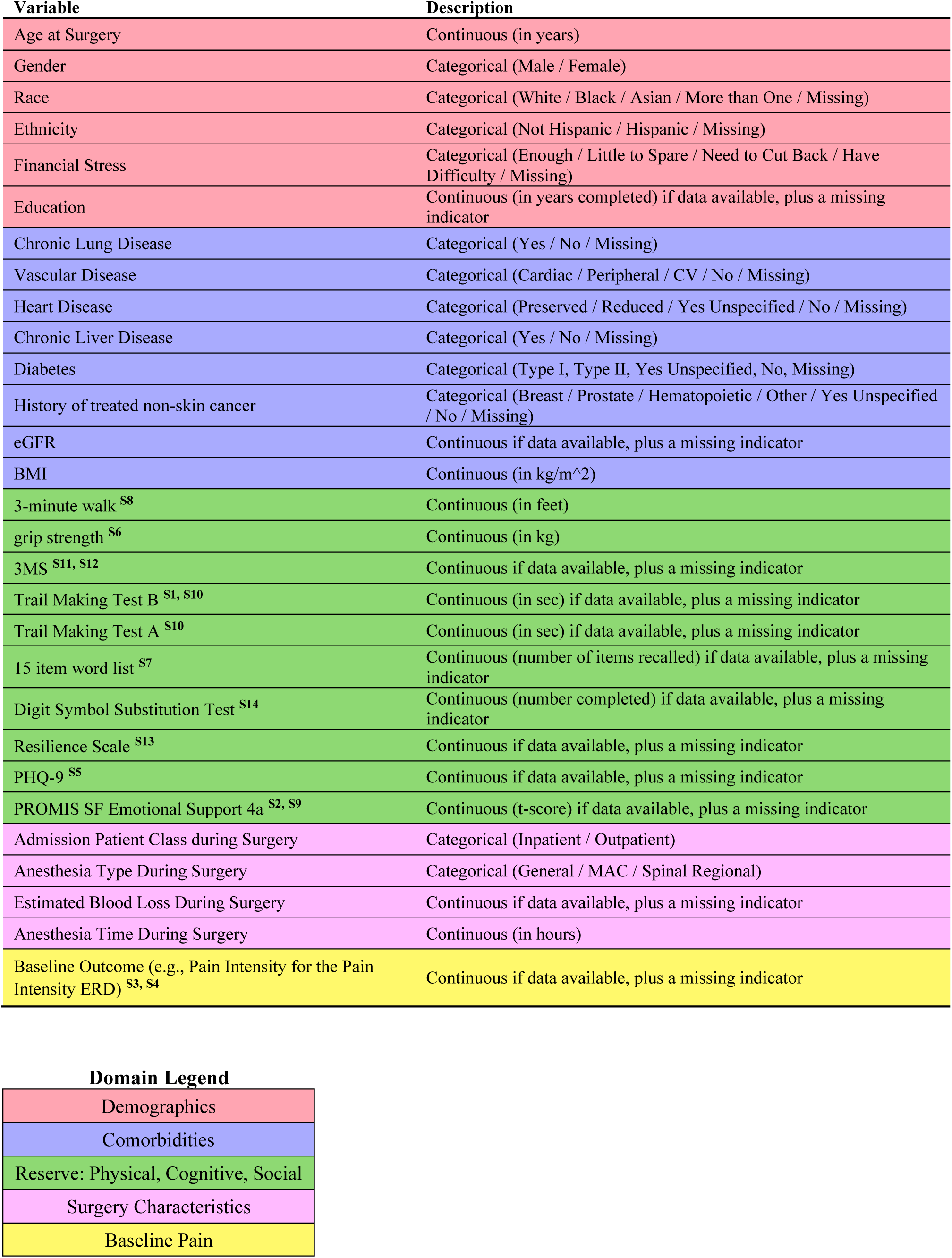
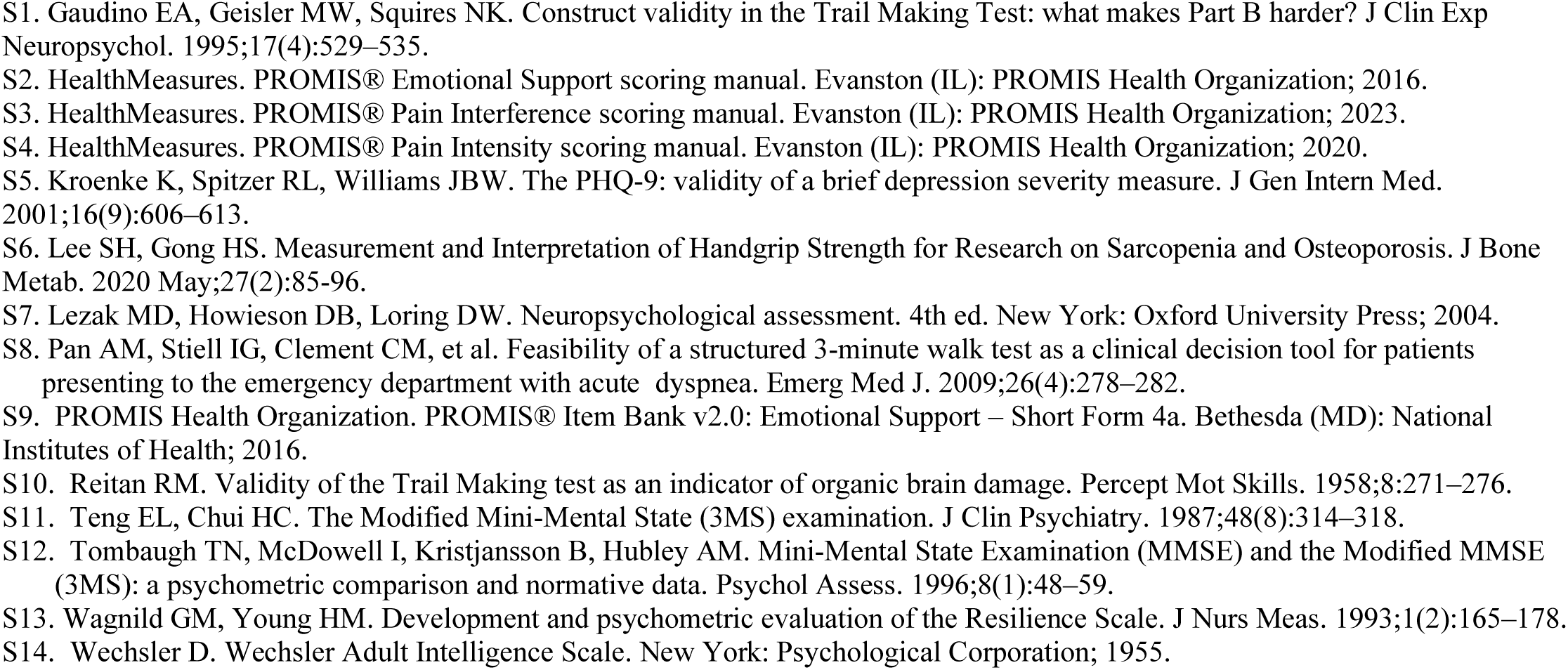
ERD Derivation Model Predictors.

**Supplemental Table 2.** Assay Kit Descriptions and Precision.

| Assay | Analyte | Median LLOD (pg/ml) | Mean Intra-assay CV (%) | Mean Inter-assay CV (%) |
| --- | --- | --- | --- | --- |
| **UPLEX Custom Proinflammatory 10-plex (MSD Cat#K151AEM-2)**<br>2-fold dilution required<br>25µl volume required<br>pg/ml final concentration units | IFN-γ | 0.766 | 1.9 | 10.1 |
|  | IL-1β | 0.084 | 4.2 | 3.1* |
|  | IL-2 | 0.579 | 10.5 | 7.5* |
|  | IL-4 | 0.01 | 7.5 | 0.9* |
|  | IL-5 | 0.098 | 2.5 | 38.4 |
|  | IL-6 | 0.152 | 3.4 | 22.9 |
|  | IL-8 | 0.094 | 2.9 | 10.3 |
|  | IL-10 | 0.047 | 2.8 | 19.3 |
|  | IL-12p70 | 0.142 | 4.4 | 5.5* |
|  | IL-13 | 0.561 | 9.2 | 5.6* |
| **UPLEX Custom Cytokine 10-plex (MSD Cat#K151AEM-2)**<br>2-fold dilution required<br>25µl volume required<br>pg/ml final concentration units | TNF-α | 0.184 | 3.6 | 37.9 |
|  | Eotaxin | 3.47 | 8.7 | 3.7 |
|  | IP-10 | 0.086 | 1.2 | 7.2 |
|  | MCP-1 | 1.94 | 7 | 8.7 |
|  | MIP-1α | 1.85 | 4.1 | 24 |
|  | MIP-1β | 0.124 | 2.1 | 10.2 |
|  | IFNα2a | 0.644 | 8.7 | 3.5* |
|  | IL-7 | 0.264 | 6.2 | 6.9* |
|  | IL-15 | 0.053 | 2 | 10.5 |
|  | IL-18 | 0.676 | 2.6 | 4.1 |
| **Custom UPLEX/RPLEX 10-plex (MSD Cat#K151AEM-2)**<br>2-fold dilution required<br>25µl volume required<br>pg/ml final concentration units | sCD30 | 158 | 9 | 27.5 |
|  | GROα | 0.028 | 8 | 5.7 |
|  | Granzyme B | 0.336 | 2.6 | 8.3 |
|  | I-TAC | 0.14 | 3.3 | 11.7 |
|  | IL-1Ra | 0.558 | 10.1 | 4.8 |
|  | IL-27 | 1.65 | 3.6 | 10.1 |
|  | IL-37 | 0.676 | 9.5 | 11.2* |
|  | Leptin | 7.93 | 6.2 | 9.1 |
|  | MIG | 0.296 | 2.2 | 5.5 |
|  | TRAIL | 0.266 | 1.7 | 16.7 |
| **RPLEX GDF-15 (MSD Cat#K151YDR)**<br>100-fold dilution required<br>10µl volume required<br>pg/ml final concentration units | GDF-15 | 0.122 | 3.7 | 5.2 |
| **Custom RPLEX 2-plex LIF/TGF-α (MSD)**<br>undiluted<br>25µl volume required<br>pg/ml final concentration units | LIF | 1.45 | 4.3 | 13 |
|  | TGF-α | 0.13 | 1.8 | 2.8 |
| **Custom UPLEX 2-plex gp130/TNFR1 (MSD)**<br>100-fold dilution required<br>5µl volume required<br>pg/ml final concentration units | gp130 | 5.49 | 1.6 | 4.7 |
|  | TNFR1 | 0.06 | 1.56 | 4.65 |
| **RPLEX Human LBP (MSD Cat#K151K5R)**<br>500-fold dilution required<br>5µl volume required<br>pg/ml final concentration units | LBP | 110 | 4.6 | 6.3 |
| **Serpine E1/PAI-1 (R&D Cat#DSE100)**<br>2-fold dilution required<br>25µl volume required<br>ng/ml final concentration units | PAI-1 | 0.059 | 5.2 | 5.4 |
| **Human Activin A (R&D Cat#DAC00B)**<br>5-fold dilution required<br>20µl volume required<br>pg/ml final concentration units | Activin A | 3.67 | 3.9 | 6.2 |
| **VPLEX Vascular Injury (MSD Cat#K15198D)**<br>1000-fold dilution required<br>10µl volume required<br>ng/ml final concentration units | SAA | 21.2 | 3.8 | 11.3 |
|  | CRP | 3 | 2.1 | 6.7 |
|  | VCAM-1 | 11 | 6.2 | 7.1 |
|  | ICAM-1 | 2.05 | 3.8 | 9.9 |
| **UPLEX TGF-β (MSD Cat#K151XWK)**<br>1.34-fold dilution required<br>25μl volume required<br>pg/ml final concentration units | TGF-β | 28.3 | 11.1 | 9.8 |
| **CMV IgG (Gold Standard Diagnostics Cat#720-320)**<br>101-fold dilution required<br>10μl volume required<br>EU/ml final concentration units | CMV IgG | 10 EU/ml | 5.0 | 4.8 |
| **Cortisol (Monobind Cat#3625-300)**<br>undiluted<br>25μl volume required<br>μg/dL final concentration units | Cortisol | 0.34 μg/dL | 8.5 | 6.7 |
| **D-dimer (Biomedica Diagnostics Cat#602)**<br>21-fold dilution required<br>25μl volume required<br>ng/ml final concentration units | D-dimer | 2-4 ng/ml | 3.7 | 6.9 |
| **Fibrinogen (Hyphen Biomed Cat#RK024A)**<br>200,000-fold dilution required<br>5μl volume required<br>ng/ml final concentration units | Fibrinogen | 2.5 ng/ml | 3.7 | 7.2 |
| **LPS (Biovendor Cat#890030)**<br>40-fold dilution required<br>10μl volume required<br>EU/ml final concentration units | LPS | 0.005 EU/ml | 3.5 | 7.3 |

## Notes

### Competing Interest Statement

The authors have declared no competing interest.

### Clinical Protocols

https://pubmed.ncbi.nlm.nih.gov/34325481/

### Author Declarations

IRB of Duke University gave ethical approval for this work.

### Summary of Updates

Standardize molecular symbols, fix grammatical and typographical errors, modify results and discussion to improve clarity.

